# Targeting PTP1B and DUSP4 phosphatases to Boost Tregs: A Novel Therapy for Polyendocrine Metabolic Ovarian Syndrome (PMOS) Immune Dysfunction

**DOI:** 10.64898/2026.05.30.26354518

**Authors:** Lipika Priyadarsini Patra, Betcy Susan Johnson, Krishna Pillai Jayakrishnan, Sathy M Pillai, Malini Laloraya

## Abstract

Polyendocrine metabolic ovarian syndrome (PMOS), previously called polycystic ovary syndrome (PCOS) - the most common reproductive endocrinopathy in women of reproductive age, is frequently associated with chronic low-grade inflammation and immune dysregulation. Beyond hyperandrogenism and ovulatory dysfunction, women with PMOS exhibit reduced regulatory T cell (Treg) levels and impaired STAT5 phosphorylation. This study investigates the molecular basis of the defective STAT5 signalling in PMOS. No significant difference in plasma IL2 levels is observed in PMOS women versus normal subjects. Analysis of 102 PMOS patients and 102 controls reveals significantly decreased *JAK2* expression alongside increased expression and activity of the phosphatases *PTP1B* (Protein Tyrosine Phosphatase 1B), *TCPTP* (T cell Protein Tyrosine Phosphatase), and *DUSP4* (Dual Specificity Protein Phosphatase), in leukocytes of PMOS women. In isolated Tregs, only *PTP1B* and *DUSP4* were significantly upregulated. DUSP4 expression positively correlates with serum testosterone and luteinizing hormone levels, linking hormonal imbalance with immune defects. Functional experiments show that silencing PTP1B and DUSP4 enhances IL2-induced Treg generation. Our collective findings identify phosphatase-mediated inhibition of STAT5 signalling as a key mechanism underlying Treg deficiency in PMOS and highlight PTP1B and DUSP4 as potential therapeutic targets to restore immune tolerance and improve reproductive outcomes.

**Graphical Abstract:** Summary of our findings of overexpressed phosphatases as negative regulators of IL-2 driven Treg expansion in PMOS suggesting them to be druggable targets.

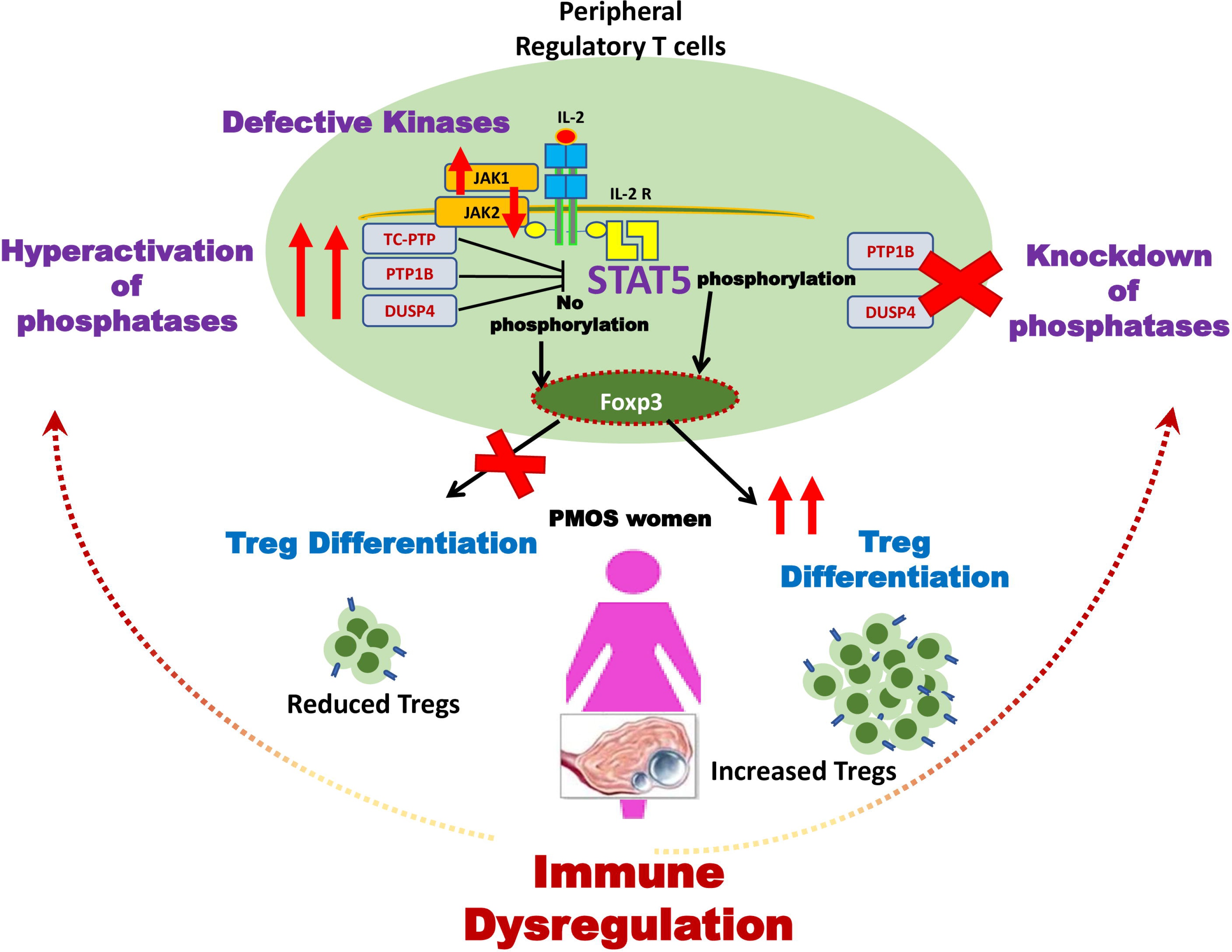

## INTRODUCTION

Polyendocrine metabolic ovarian syndrome (PMOS), formerly called polycystic ovary syndrome (PCOS), is a leading multifactorial disease affecting a notable proportion of fertile women. According to a review study, it affects approximately 4–21% of women globally, depending on various diagnostic criteria ^1^. Based on World Health Organisation (WHO) reports, PMOS affects nearly 6-13% of fertile female (https://www.who.int/news-room/fact-sheets/detail/polycystic-ovary-syndrome). It is associated with heterogeneous clinical manifestations including irregular menstrual cycles, oligo/anovulation, hyperandrogenism ^2^. Additionally, it is linked to a wider range of co-morbidities, such as ovulatory dysfunction, elevated serum LH, elevated LH/FSH ratio ^3^, insulin resistance (IR) ^4^, obesity, and cardiovascular disease risk. Increasing pieces of evidences over the last decade on PMOS revealed that hyperandrogenism, obesity, and abnormal hormonal-metabolic complications of PMOS is associated with a pro-inflammatory environment ^5^. PMOS has been linked with chronic low-grade inflammation^6^ which is exacerbated during pregnancy, leading to adverse pregnancy outcome^7^. Interestingly, healthy first-degree relatives of PMOS patients also exhibit low-grade inflammation^8^. The low-grade inflammation in PMOS has been attributed to increased leukocytes^9–11^, peripheral blood inflammatory immune cells and pro-inflammatory cytokine levels^12–14^.

The mechanism of conserving the integrity of “self” by eliminating “non-self” biological materials evolved tremendously, with the role of thymus in T-cell differentiation taking a central stage^15^ and the autoimmune regulator gene as the maestro of self-antigen expression^16^. Thymic ablation and resultant lack of follicular cyst generation and anovulation under estrogen suggested T-regulatory cells depletion to be critical in cyst generation - an important characteristic of PMOS pathogenesis^17^. IL-2 receptor alpha-chain (CD25) also has a special place in autoimmunity, as it was reported that, in athymic nude mice, inoculation of CD4+ CD25-cells after depleting CD25+ cells triggered autoimmune disorders, which could be restored by the addition of CD4^+^CD25^+^ cells^18^. Later, the work from our lab reported a dynamic preovulatory reduction of the CD4^+^CD25^+^CD127^low^ T regulatory cells (Treg) in PMOS women compared to normal women and suggested a defective immune tolerance in PMOS women, thereby explaining increased implantation failures and abortions in them^19^.

Tregs are vital for maintaining the immune homeostasis and their imbalance can tilt it from autoimmunity to immunodeficiency and infection susceptibility^20^. Tregs are subtypes of CD4^+^, and interleukin 2 (IL-2) receptor α chain^+^ (CD25^+^) T cells, and their differentiation as well as functional activity depends principally on the expression of FOXP3 (Forkhead box P3)^21,22^ as well as the low abundance of CD127 (IL-7R)^23^. The Treg generation and proliferation loop involves many pathways, but the IL-2R/CD28/T cell receptor (TCR) signaling pathway plays a significant paradoxical role in Treg dynamics^24,25^. According to recent studies, IL-2 signaling is necessary for the proliferation and maintenance of an adequate number of Tregs in the peripheral pool^26,27^. IL-2 induces differentiation and expansion of Treg cells by stimulating FOXP3 expansion via another critical family of transcription factor, Signal Transducer and Activator of Transcription 5 (STAT5)^28,29^. STAT5 gets activated upon phosphorylation and subsequently translocates to the nucleus for binding with the CNS2 (Conserved Non-coding Sequence-2) position of the *FOXP3* locus, leading to the differentiation of Treg cells^30^.

The functional regulators for STAT5 activation can occur at a post-translation level by most importantly, tyrosine phosphorylation. In our previous report, Krishna et al found that exogenous IL-2 stimulation was insufficient to increase FOXP3 expression in PMOS women^19^. They discovered that although exogenous IL-2 increased activated STAT5 (pSTAT5A/B) levels in healthy women, PMOS women’s PBMC failed to show any phosphorylated STAT5B^19^. STAT5 phosphorylation depends on the net impact of phosphorylating mechanisms under control of Janus Kinases (JAK) and dephosphorylating mechanisms involving phosphatases. Previous studies showed that the activation of the cytokine-JAK-STAT signalling pathway can be negatively regulated by several phosphatases, like PTPs (Protein Tyrosine Phosphatases), including protein tyrosine phosphatase 1B (PTP1B)^31^, and T-cell protein tyrosine phosphatase (TCPTP)^32^. It was also observed that a notable over-expression of PTP1B plays a vital role in prolactin-induced STAT5 dephosphorylation^31^. STAT5 activity can also be inhibited by dual specificity protein phosphatase (DUSPs), especially DUSP4. Besides dephosphorylation, DUSP4 overexpression suppresses STAT5 transcriptional activity by destabilizing STAT5 protein^33^. In the current study, we investigate the mechanisms underlying the dephosphorylated state of STAT5 in PMOS to better understand the basis of reduced peripheral regulatory T cells in this condition.

## RESULTS

### Study group characteristics

All subjects included in our study are of similar age groups. The body mass index (BMI) of the PMOS group is considerably higher than the control (p-value <0.05). Clinical and biochemical characteristics of subjects enrolled in our study are reported in Table 1A and all values are shown as mean ± SEM(Standard Error Mean). No change in FSH level is observed between the two groups. The level of LH is notably high in PMOS patients (p-value <0.0001) as compared to the control females. Also, the PMOS patients showed significantly high LH/FSH ratio when compared to controls (p-value <0.0001). The total testosterone level (p-value<0.0001) as well as the DHT level (p-value<0.01) are significantly increased in PMOS women compared to control females. A graphical Representation is shown in Figure S1.

**Table 1A:** Clinical and biochemical parameters of the Control and PMOS groups. *The clinical and biochemical parameters of control (n=102) and PMOS (n=102) groups.* All values are presented as mean ± SEM. p-value< 0.05 is considered statistically significant.

| Characteristics | Control | PMOS | p-value |
| --- | --- | --- | --- |
| Age (years) | 28.2 ± 0.31 | 28.9 ± 0.43 | >0.05 |
| BMI (kg/m<sup>2</sup>) | 22.9 ± 0.33 | 24.1 ± 0.36 | <0.05 |
| LH (mIU/ml) | 6.1 ± 0.56 | 14.6 ± 0.33 | <0.0001 |
| FSH (mIU/ml) | 6.9 ± 0.35 | 6.15 ± 0.24 | >0.05 |
| LH/FSH | 0.92 ± 0.09 | 2.9 ± 0.15 | <0.0001 |
| Testosterone (ng/dl) | 28.5 ± 0.43 | 45.8 ± 1.81 | <0.0001 |
| DHT (ng/ml) | 0.15 ± 0.01 | 0.2 ± 0.01 | <0.01 |
| Menstrual cycle | 28-35 days | Menstrual cycle > 40 days |  |

### Phenotypic categorization of PMOS participants of the study group

PMOS individuals (n=102) included in this study were characterized into four main phenotypic subgroups: phenotype A, B, C, and D based on the diagnostic criteria of hyperandrogenism (HA), Ovulatory Dysfunction (OD), and Polycystic Ovarian Morphology (PCOM) based on the NIH (2012), an extension of ESHRE/ASRM, 2003. Our study group showed that phenotype A (HA+OD+PCOM) is the most prevalent subgroup with 55.9% of cases. While phenotype B (HA+OD) is the least found subgroup with 4.9% cases. We found that phenotype C (HA+PCOM) is the second highest phenotype with 23.5% cases and phenotype D (OD+PCOM) with 15.7% cases in the extreme southern part of India (Kerala). There is no statistically significant difference in the levels of LH, FSH, LH/FSH, and DHT among the four PMOS phenotypic populations. We observed a significantly higher level of testosterone in the population of phenotypes A, B, and C compared to phenotype D (Table 1B). Reproductive complications were observed in all four groups.

**Table 1B:** Clinical and biochemical parameters in individuals with four different PMOS phenotypes. The clinical and biochemical parameters of four PMOS phenotypes (phenotype A, B, C, and D). The number of subjects of each PMOS phenotypes in the study group are represented in percentage (%). All values are presented as mean ± SEM.

| Characteristics | Phenotype A<br>(55.9%) | Phenotype B<br>(4.9%) | Phenotype C<br>(23.5%) | Phenotype D<br>(15.7%) |
| --- | --- | --- | --- | --- |
| BMI (kg/m<sup>2</sup>) | 24.2 ± 0.47 | 21.5 ± 2.52 | 23.7 ± 0.72 | 25.4 ± 0.83 |
| LH (mIU/ml) | 14.8 ± 0.4 | 12.2 ± 0.55 | 15.3 ± 0.81 | 13.6 ± 0.81 |
| FSH (mIU/ml) | 5.9 ± 0.30 | 7.0 ± 1.64 | 6.3 ± 0.6 | 6.7 ± 0.54 |
| LH/FSH | 3 ± 0.21 | 2 ± 0.32 | 3.2 ± 0.39 | 2.2 ± 0.14 |
| Testosterone (ng/dl) | 49.2 ± 1.91 | 50.7 ± 7.23 | 54 ± 3.11 | 20.1 ± 3.49 |
| DHT (ng/ml) | 0.23 ± 0.02 | 0.17 ± 0.04 | 0.2 ± 0.02 | 0.15 ± 0.02 |
| Ovary size (cm<sup>3</sup>) | 13.37 ± 0.6 | 9.2 ± 1 | 12.1 ± 0.5 | 11.38 ± 2.8 |

### IL-2 is not the key factor for low STAT5 activity in PMOS condition

IL-2 mediated activation of STAT5 is critical for the differentiation of T regulatory cells, as binding of IL-2 with IL-2 receptor initiates STAT5 signalling. Therefore, estimation of the secretory levels of IL-2 is necessary to understand the underlying mechanism of low pSTAT5 expression and low Treg numbers in PMOS patients. To estimate the secretory levels of IL-2, we performed an ELISA and observed no significant difference in the level of IL-2 between control (n=37) and PMOS (n=37) females (Figure 1A).

**Figure 1:**
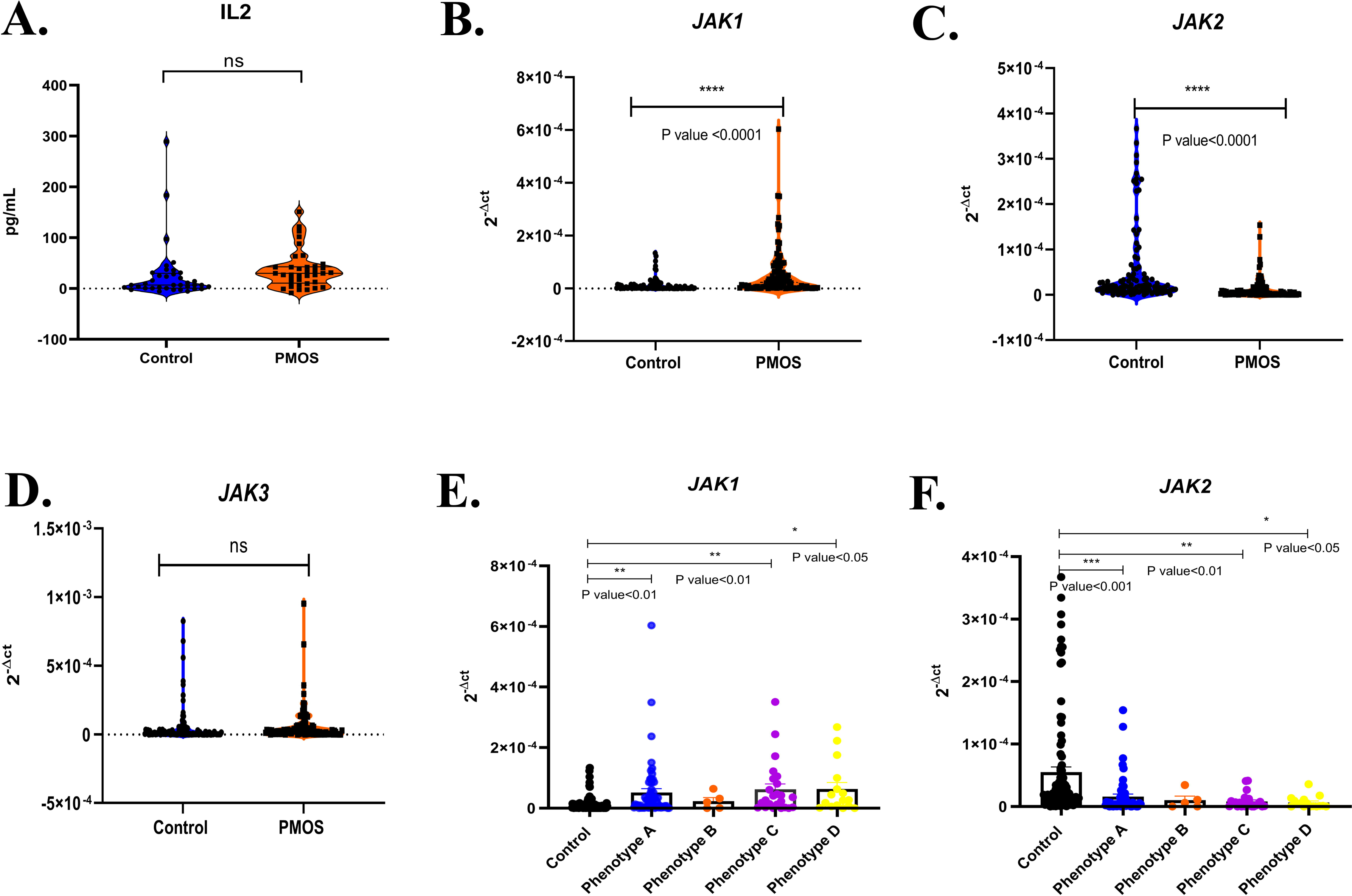
Unaltered plasma IL-2 and differential Janus Kinases expression in leukocytes of PMOS subjects and in four PMOS phenotypes. Plasma level IL-2 in control (n=37) and PMOS (n=37) subjects and altered gene expression of *JAKs* in the total leukocyte RNA of 102 control and 102 PMOS subjects. A: Dot plot graphical representation of IL-2 level in the control and PMOS subjects. B-D: Dot and violin plot graphical representation of real-time gene expression level in *JAK1, JAK2* and *JAK3*. Each dot represents an individual subject. Each reaction was performed in triplicates and the ΔCt method was used to calculate the gene expression difference between control and PMOS subjects. 2^−ΔCt^ values of each subject are plotted on the Y-axis of the graph. Unpaired student’s T-tests were performed using GraphPad Prism 8 and p-value< 0.05 were set to be significant. Graph E and F: Dot and bar graph representation of relative gene expression level of *JAK1,* and *JAK2* among the control and PMOS phenotype A, B, C, and D. The control group represented black dots, phenotype A in blue dots, phenotype B in orange dots, phenotype C in violet dots and phenotype D in yellow dots. One-way ANOVA was performed using GraphPad Prism 8 and p-value< 0.05 were set to be statistically significant.

### Altered gene expression of Janus Kinases in leukocytes of PMOS patients

The functional regulation for STAT5 activation initiates with the binding of IL-2 with its high-affinity IL-2 receptor (IL-2R) which is associated with the cytoplasmic Janus Kinase (JAK) activity. After auto-phosphorylation of JAK, the STAT5 gets phosphorylated and translocates to the nucleus to activate FOXP3(Forkhead box P3). Therefore, the expression of JAKs is crucial to the STAT5 phosphorylation and signalling. Upon qRT-PCR, the relative mRNA expression analysis of Janus kinases in leukocytes of control (n=102) and PMOS (n=102) subjects revealed an upregulation of JAK1 in PMOS patients (FC=2.19, p-value < 0.0001) compared to control women (Figure 1B). While the transcript levels of JAK2 showed down-regulation in PMOS individuals (FC= −2.15, p-value < 0.0001) (Figure 1C). The relative expression of JAK3 between both the control and PMOS groups showed no significant change (Figure 1D).

### Relative gene expression of *JAK1* and *JAK2* in four PMOS phenotypes

The relative gene expression levels of *JAK1* and *JAK2* in the leukocytes of four PMOS phenotypes were also analysed to understand the phenotypic variations of Janus Kinase expression among our PMOS group. The transcript levels of *JAK1* showed a significant elevation in phenotype A (HA+OD+PCOM), phenotype C (PCOM+HA), and phenotype D (OD+PCOM) as compared to the control groups (Figure 1E). Whereas the relative expression of *JAK2* significantly reduced in phenotype A, phenotype C and phenotype D when compared to the control subjects (Figure 1F).

### Upregulation of phosphatases of the pSTAT5 pathway in leukocytes of PMOS patients

Phosphatases such as PTP1B, TCPTP, and DUSP4 are crucial components of the negative loop that down-regulates STAT5 activity by causing its dephosphorylation. Upon qRT-PCR analysis in the leukocytes of control (n=102) and PMOS (n=102) subjects, the PMOS patients showed a significant upregulation in the relative gene expression of *PTP1B* (FC=1.02, p-value<0.01), *TCPTP* (FC=0.86, p-value<0.01), and *DUSP4* (FC=2.2, p-value<0.0001) compared to control subjects, as shown in Figure 2A-2C. We further assessed the relative gene expression levels of *PTP1B, TCPTP*, and *DUSP4* in each PMOS individual to understand the subject-level variations among our PMOS group and this is shown in Figure 2D. The variations in the gene expression level of these phosphatases showed a different expression pattern in each PMOS subject, indicating that not all phosphatases are expressed equally in each subject. Notably, DUSP4 shows the highest expression in several subjects.

**Figure 2:**
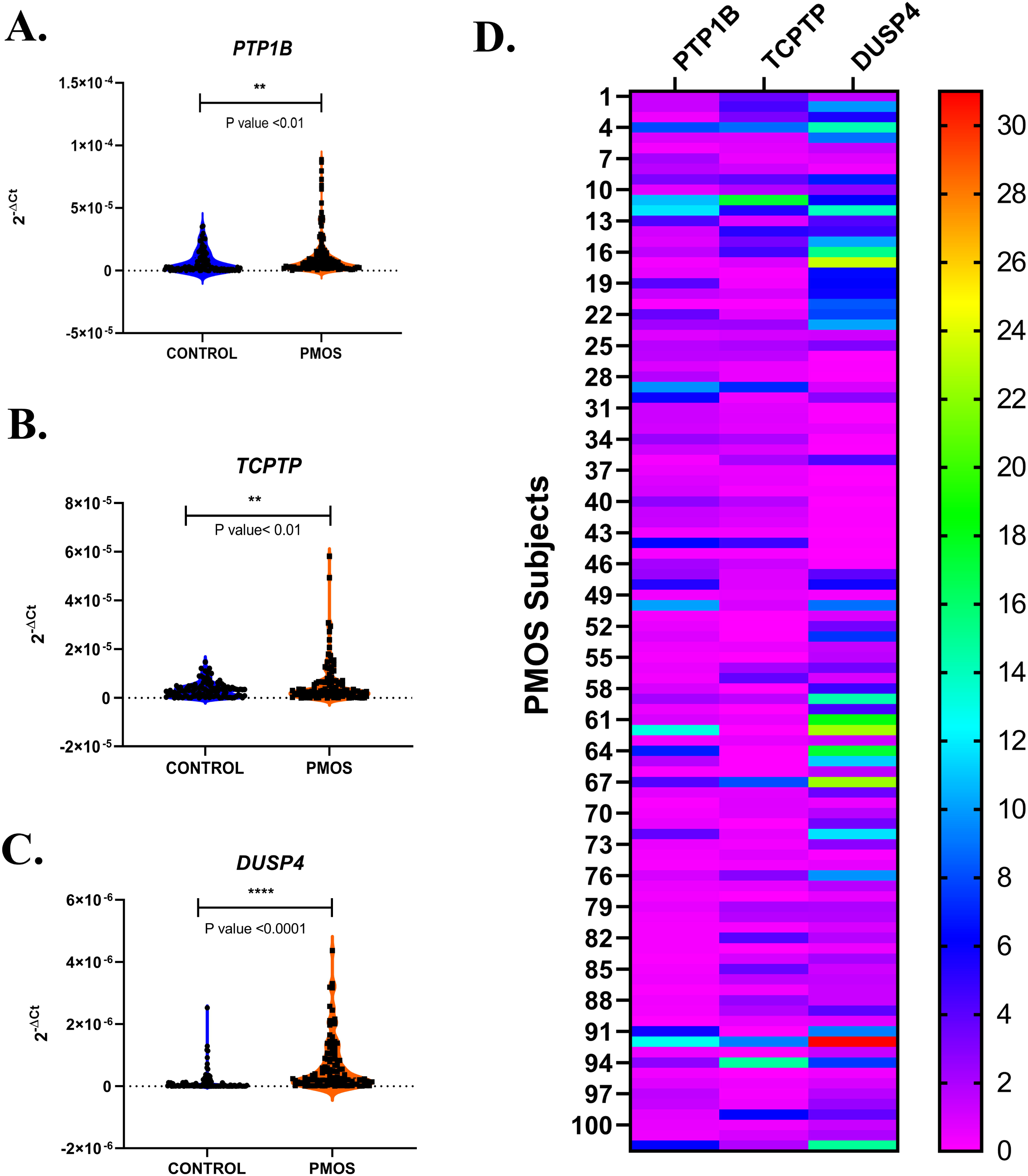
Elevated gene expression of phosphatases in leukocytes of PMOS subjects. Increased phosphatase gene expression in the total leukocyte RNA of 102 control and 102 PMOS subjects. A-C: Dot and violin plot graphical representation of real-time gene expression level in *PTP1B, TCPTP*, and *DUSP4*. Each dot inside the violin plot represents an individual subject. Each reaction was performed in triplicates and the ΔCt method was used to calculate the gene expression difference between control and PMOS subjects. 2^−ΔCt^ values of each subject are plotted on the Y-axis of the graph. Unpaired student’s T-tests were performed using GraphPad Prism 8 and p-value< 0.05 were set to be significant. Graph D: Heatmap analysis showing the expression level of phosphatases among each PMOS subject (n=102). The rainbow colour gradation shows the expression level in individual subjects with pink colour (lowest value) and red colour (highest value).

### Relative gene expression of *PTP1B, TCPTP,* and *DUSP4* in four PMOS phenotypes

We also explored the relative gene expression levels of *PTP1B, TCPTP,* and *DUSP4* in the leukocytes of four PMOS phenotypes to understand the phenotypic variations among our PMOS group. The transcript levels of *PTP1B, TCPTP*, and *DUSP4* showed a significant alternation in phenotype A (HA+OD+PCOM) compared to control groups. *DUSP4* also showed a significant upregulation in phenotype C (PCOM+HA) as compared to the control subjects (Figure 3A-3C). The expression pattern of *PTP1B, TCPTP,* and *DUSP4* among the four PMOS phenotypes was also analysed and shown via Heat-map in Figure 3D-3G. All three phosphatases: *PTP1B, TCPTP,* and *DUSP4* were expressed in major number of individuals of phenotype A, 56%, 47% and 77% respectively, with DUSP4 showing the greatest increase (Figure 3D). Whereas 40% individuals of phenotype B and C showed increased expression of *PTP1B and DUSP4* compared to enhanced *TCPTP* in 20% individuals(Figure 3 E and F). 50% individuals of phenotype D had higher gene expression level of *DUSP4* while only 31% subjects showed elevated *PTP1B* and *TCPTP* (Figure 3G), although the change in DUSP4 in Phenotype D is not significant. This signifies that *DUSP4* appears to be the critical phosphatase that is significantly upregulated in PMOS phenotypes A & C. Finally, we observe that each subject expresses one or the other phosphatase, nonetheless the phosphatases are not clustered together specifically for a particular phenotype.

**Figure 3:**
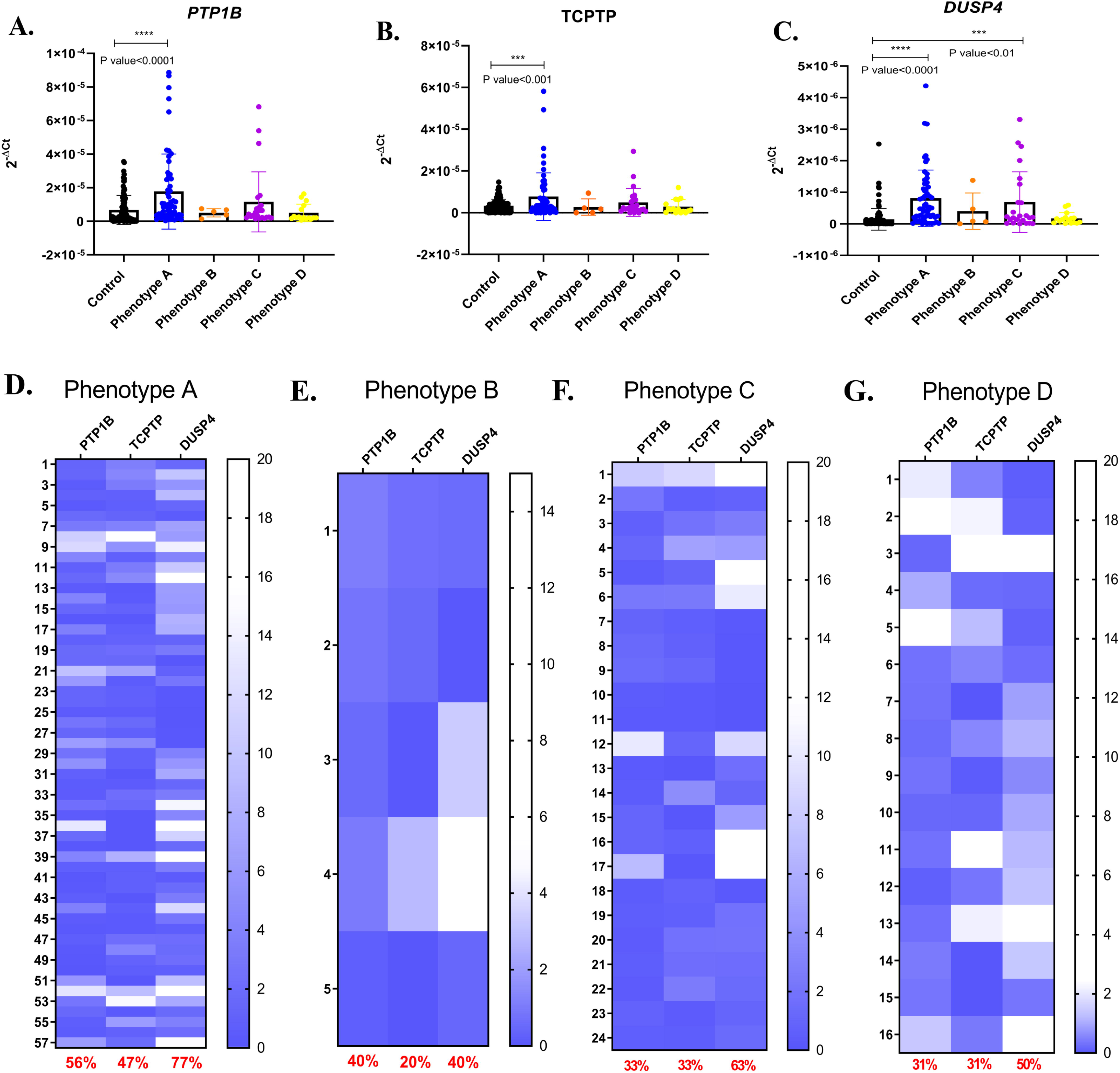
Phosphatases gene showed expression variation in different PMOS phenotypes. Detailed PMOS subtype (phenotype A, B, C, and D) and each subject analysis of relative mRNA expression of phosphatases in 102 control and 102 PMOS subjects. Graph A-C: Dot and bar graph representation of relative gene expression level of *PTP1B, TCPTP*, and *DUSP4* among the control and PMOS phenotype A, B, C, and D. The control group represented black dots, phenotype A in blue dots, phenotype B in orange dots, phenotype C in violet dots and phenotype D in yellow dots. Each dot inside the bar graph represents an individual subject. 2^−ΔCt^ values of each subject are plotted on the Y-axis of the graph. One-way ANOVA was performed using GraphPad Prism 8 and p-value< 0.05 were set to be statistically significant. Graph D-G: Heatmap analysis showing the expression level of phosphatases gene expression among the PMOS subjects of four different phenotypes (Phenotype A, B, C and D respectively). The blue colour gradation shows the expression level in individual subjects with blue colour (lowest value) and white colour (highest value).

### DUSP4 in PMOS patients showed correlation with hyperandrogenism and higher LH level

The correlation between the three phosphatases with hyperandrogenism (since high testosterone is the hallmark of PMOS), and other biochemical parameters, was determined by performing a linear regression analysis between these factors. Our analysis revealed a significant correlation between the gene level expression of *DUSP4* with LH (p-value < 0.01) (Figures 4K) and *DUSP4* with testosterone (p-value < 0.05) (Figures 4M), thereby explaining its absence in Phenotype D, which is devoid of hyperandrogenemia. Interestingly, *PTP1B* and *TCPTP* did not show any significant correlation with the elevated parameters such as LH, LH/FSH, testosterone, DHT, and BMI (Figure 4A-4E, and 4F-4J respectively), suggesting a potent role of DUSP4 in PMOS pathogenesis.

**Figure 4:**
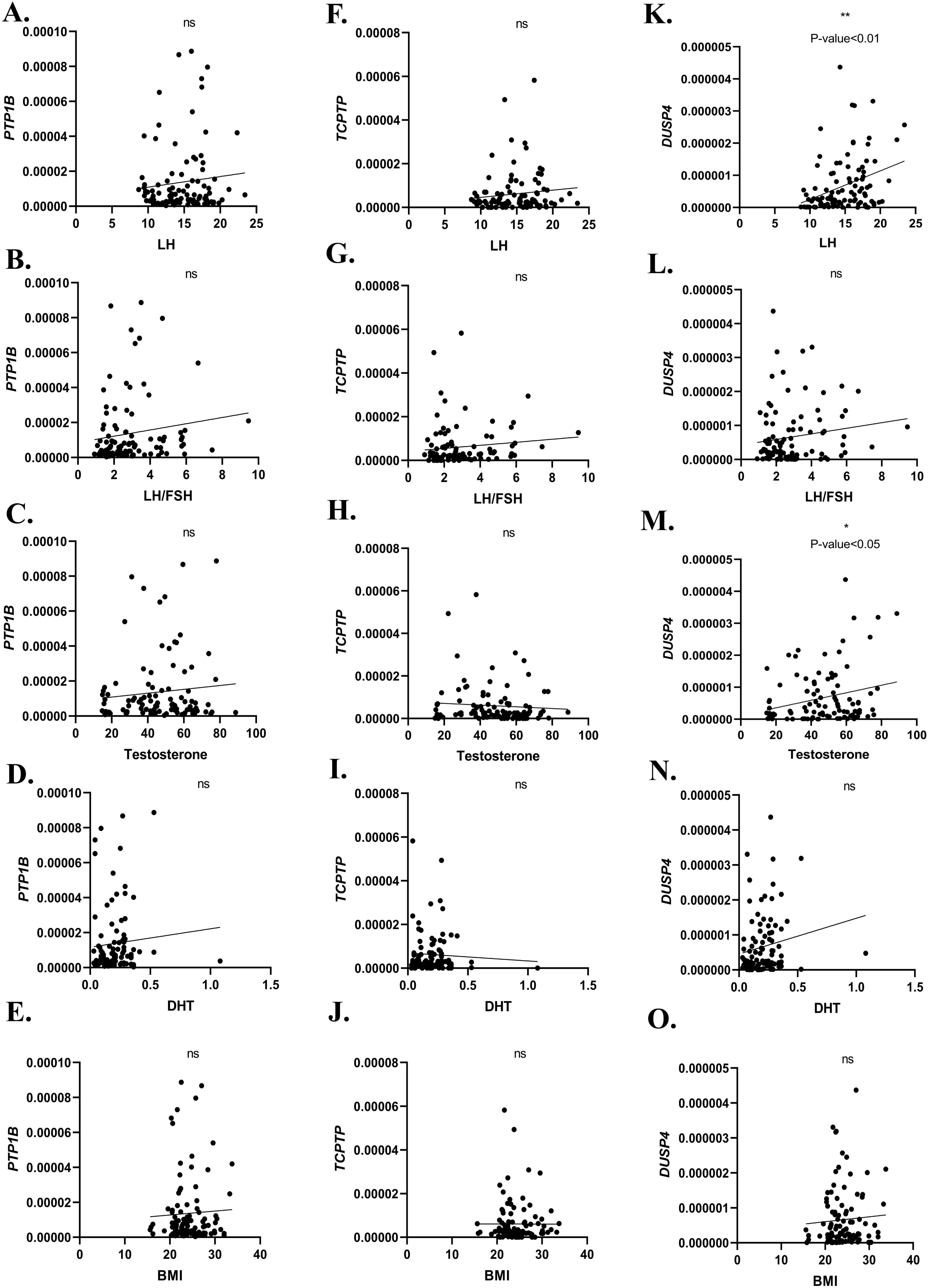
Regression correlation analysis of phosphatases with all biochemical parameters. Graphs (A-E) show the linear regression correlation analysis between *PTP1B* with LH, LH/FSH, Testosterone, DHT, and BMI. Graphs (F-J): Linear regression correlation analysis between *TCPTP* with LH, LH/FSH, Testosterone, DHT, and BMI. Graphs (K-O) Linear regression correlation analysis between *DUSP4* with LH, LH/FSH, Testosterone, DHT, and BMI. P-values <0.05 are considered statistically significant.

### PBMCs of PMOS patients showed increased phosphatase activity

To understand the enzymic activity of phosphatases in the PMOS condition, we measured the enzymatic capability in dephosphorylating proteins from tyrosine phosphatases using a fluorometric phosphatase assay kit in the PBMCs of control (n=20) and PMOS (n=23) subjects. The fluorescence reading shows a significant elevation of phosphate picomoles cleaved from the substrate (DiFMUP) in the PMOS group compared to the control. This observation signified that the significantly higher level of phosphatases also translates to increased phosphatase activity in PMOS condition when compared to the control (p-value<0.001) as shown Figure 5A. To revalidate, the expression levels of *PTP1B, TCPTP*, and *DUSP4* in the subjects in which phosphatase activity was assessed, qRT-PCR was performed. Again, we observed significantly higher expression of *PTP1B* (FC=2.49, p-value<0.05), TCPTP (FC=1.01, p-value<0.05), and *DUSP4* (FC=2.7, p-value<0.05) in these PMOS subjects (Figure 5B-5D), reiterating our previous observations. Our findings confirmed a significant up-regulation of phosphatase activity in the PBMCs of the PMOS subjects.

**Figure 5:**
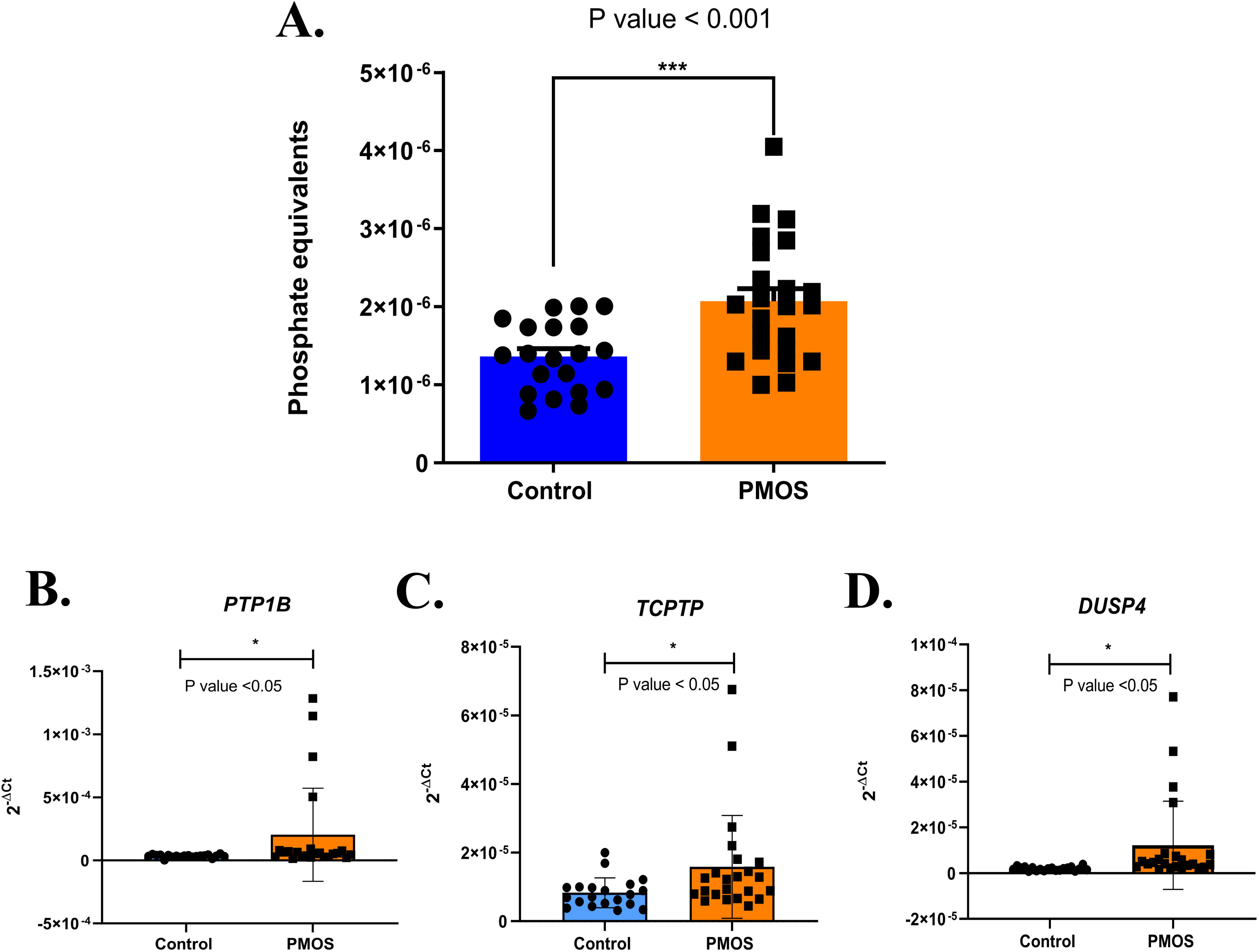
Elevated Phosphatase activity in PBMCs of PMOS subjects compared to control groups. Increased phosphatase activity in the PBMCs of 20 control and 23 PMOS subjects. A: Dot and bar graph representation of phosphate equivalents released by the activity of phosphatases in the control and PMOS groups. The control group is represented in a blue colour bar with black dots and the PMOS group in an orange colour bar with black dots. B-D: Real-time gene expression analysis of *PTP1B, TCPTP*, and *DUSP4* in the subjects of phosphatase assay. Each reaction was performed in triplicates and the ΔCt method was used to calculate the gene expression difference between control and PMOS subjects. 2^−ΔCt^ values of each subject are plotted on the Y-axis of the graph. Unpaired student’s T-tests were performed using GraphPad Prism 8 and p-value< 0.05 are considered as statistically significant.

### Magnetically separated CD4^+ve^ CD25^+ve^ Treg cells revealed an up-regulation of *PTP1B,* and *DUSP4* gene expression in PMOS condition

We evaluated the expression level of *PTP1B, TCPTP*, and *DUSP4* in the magnetically isolated CD4^+ve^ CD25^+ve^ Tregs as PBMCs contain not only lymphocytes, but also monocytes, and dendritic cells. Tregs are T cell subtypes that consist of 1% - 4% of the total PBMC population, and 5% −10% of total CD4^+ve^ T cells. We have therefore carried out qRT-PCR of PTP1B, TCPTP, and DUSP4 in magnetically isolated CD4^−ve^ cells, CD4^+ve^CD25^−ve^ T effector cells, and CD4^+ve^CD25^+ve^ Treg cells to further checkwhether Tregs have higher phosphatase gene expression. The accuracy of the magnetically separated CD4^+ve^CD25^+ve^ Treg cells were validated by performing FACS as shown in Figure 6A. Our FACS analysis confirmed that 97.8% of lymphocyte (P1) magnetically separated cells were CD3-APC-H7^+ve^ T cell (P2) populations. Further, an accuracy of 97.9% of P2 populations were CD4-Per CP cy5.5^+ve^ CD25-PE^+ve^ Treg cells, as shown in Figure 6A. Hence, from this analysis it was confirmed that all the magnetically separated cells were CD4^+ve^CD25^+ve^ Treg cells. Furthermore, the transcript level expression of *PTP1B, TCPTP*, and *DUSP4* in the CD4^−ve^ cells (Figure 6B-6D) and in CD4^+ve^ CD25^−ve^ T effector cells (Figure 6E-6G) did not show any significant difference between the control (n=11) and PMOS subjects (n=13). There is a significant up-regulation in the gene expression of *PTP1B* (FC=6.55, p-value<0.05), and *DUSP4* (FC=4.62, p-value<0.05) in the CD4^+ve^ CD25^+ve^ Treg cells of PMOS women when compared to control subjects. The gene expression of *TCPTP* is not significantly altered in CD4^+ve^ CD25^+ve^ Treg cells of PMOS subjects (Figure 6H-6J). This observation illustrates an upregulation of *PTP1B* and *DUSP4* phosphatases explicitlyin the CD4^+ve^ CD25^+ve^ Treg cells in PMOS condition.

**Figure 6:**
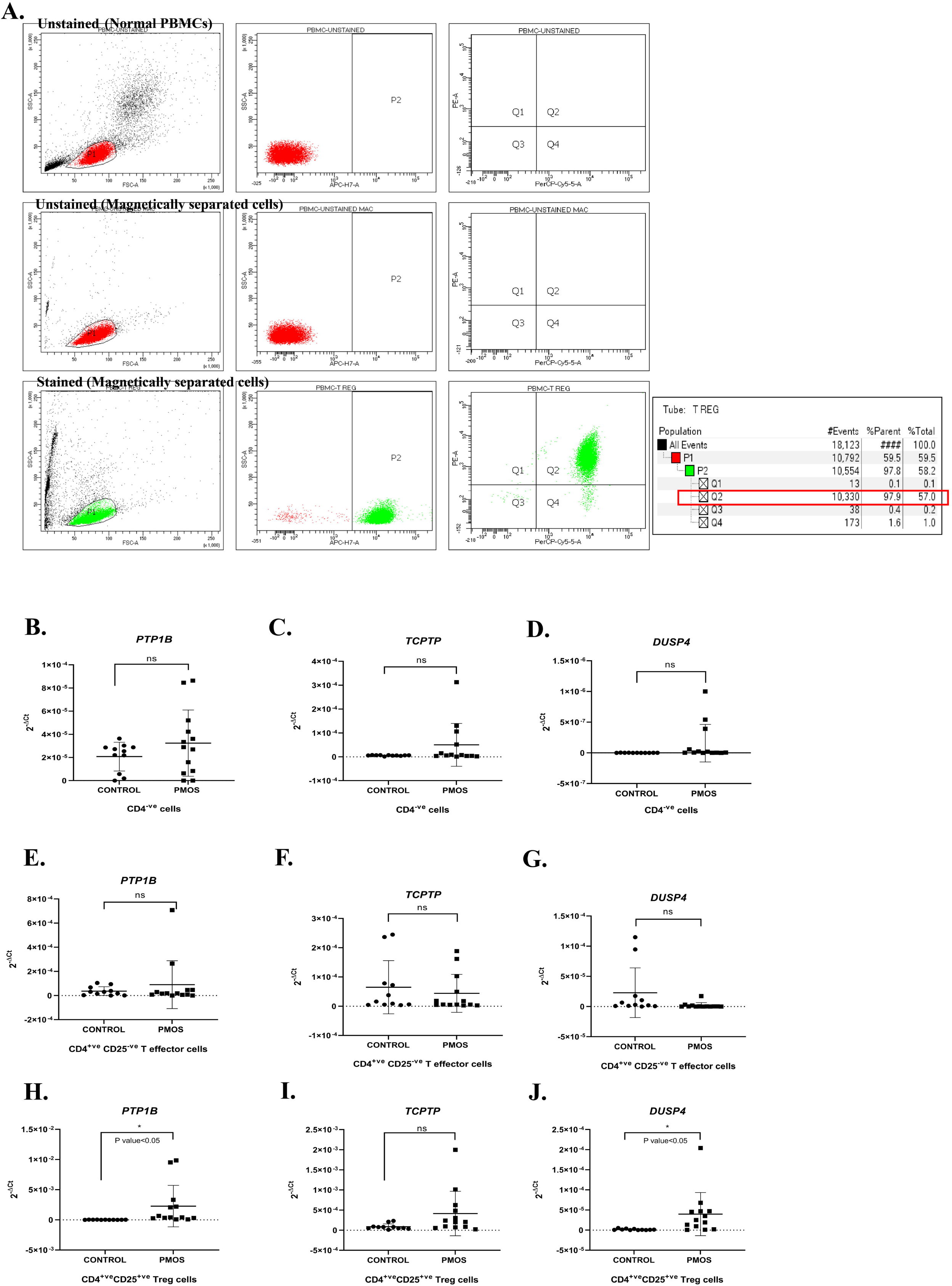
Increased gene expression of *PTP1B* and *DUSP4* in isolated CD4^+ve^ CD25^+ve^ Treg cells of PMOS subjects compared to control groups. The relative gene expression analysis of *PTP1B, TCPTP*, and *DUSP4* in the magnetically isolated CD4^−ve^ cells, CD4^+ve^ CD25^−ve^ T effector cells, and CD4^+ve^ CD25^+ve^ Treg cells of control (n=11) and PMOS (n=13) subjects. Graph A: shows the validation of magnetically isolated CD4^+ve^ CD25^+ve^ Treg cells via FACS analysis (n=01). The dot plots represent the population with P1 shows lymphocyte population, P2 shows CD3-APC-H7^+ve^ population, and Q2 shows CD4-PerCP cy5.5^+ve^ and CD25-PE^+ve^ Treg population. Gating is based on histogram of unstained and stained population (APC-H7, PE and CD4-PerCP cy5.5 respectively). Each dot represents an individual subject. The Q2 (CD4-PerCP cy5.5^+ve^ and CD25-PE^+ve^) Treg population shows an accuracy of 97.9 % positivity from the parent population (P1). B-D: Dot plot representation of the expression of phosphatases in CD4^−ve^ cells. E-G: Dot plot representation of the expression of phosphatases in CD4^+ve^ CD25^−ve^ T effector cells. H-J: Dot plot representation of the expression of phosphatases in CD4^+ve^ CD25^+ve^ Treg cells. Each dot represents an individual subject. Each reaction was performed in triplicates and the ΔCt method was used to calculate the gene expression difference between control and PMOS subjects. 2^−ΔCt^ values of each subject are plotted on the Y-axis of the graph. Unpaired student’s T-tests were performed using GraphPad Prism 8 and a p-value<0.05 are considered as statistically significant.

### CD4^+ve^ CD25^+ve^ CD127^low/-^ Treg population increased after PTP1B/DUSP4 silencing with exogenous IL-2 stimulation in PBMCs of PMOS patients

To validate the role of phosphatases (PTP1B, and DUSP4) in Treg differentiation, 12 PMOS patients underwent siRNA-based silencing of PTP1B, and DUSP4 followed by 4 days of IL-2 treatment. The efficacy of silencing is shown in Figure 7A. After individual silencing, the gene expression analysis showed a reduction of 94.8% *PTP1B* expression compared to control silencing. Similarly, the silenced PBMCs showed a reduction of 82.1% *DUSP4* gene expression. Additionally, in double silencing, the gene expression of *PTP1B* showed 97% reduction whereas, the gene expression of *DUSP4* showed 99.2% reduction. In FACS analysis, the dot plot representation of the gating of the unstained population is shown in Figure 7B. In Figure 7C, the histogram plot showed the clear gating of negative and positive populations in unstained and stained cells. The effect of *PTP1B* and *DUSP4* silencing on Treg differentiation was assessed by FACS analysis detailed below:

**Figure 7:**
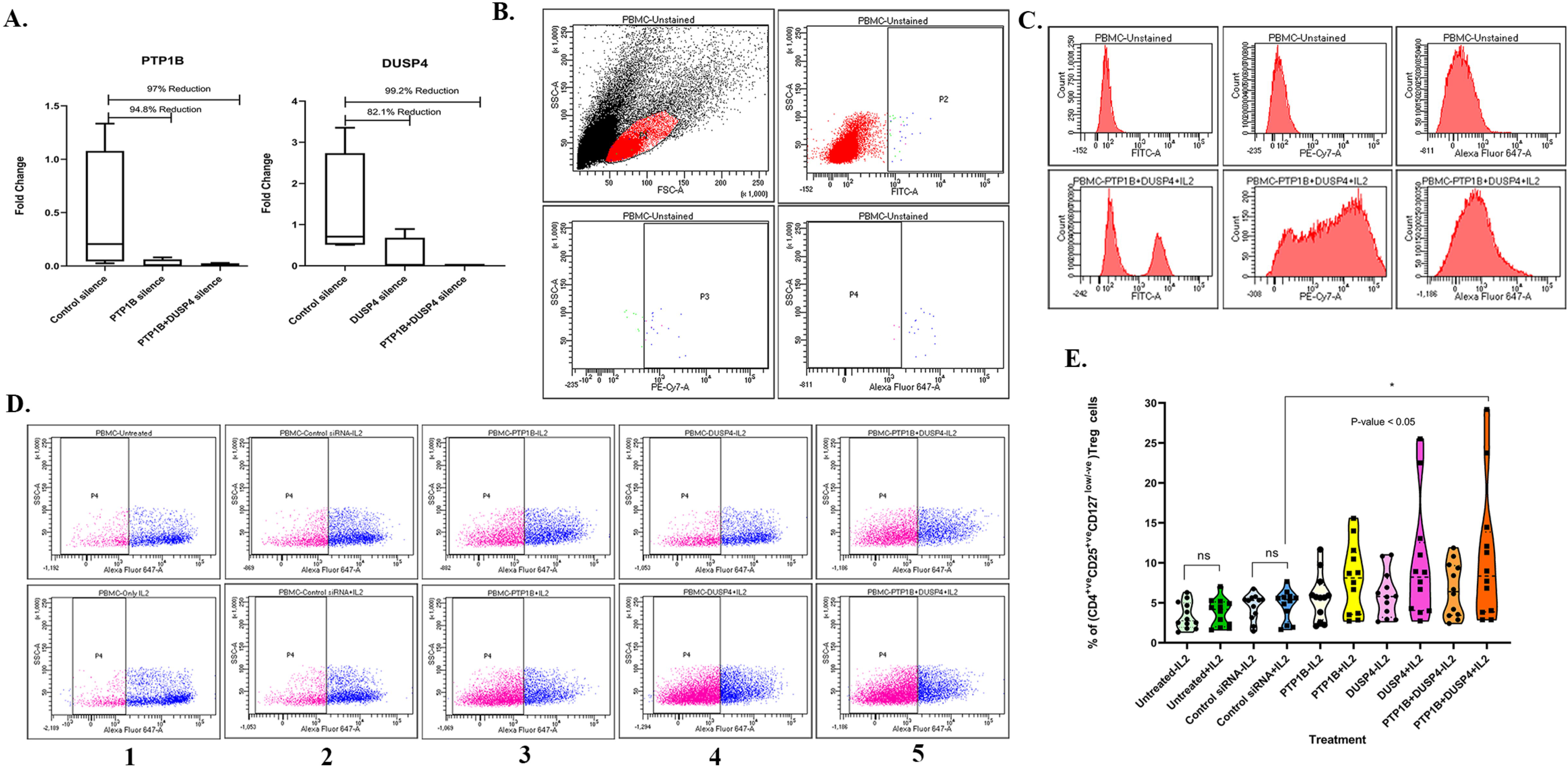
Elevation of CD4^+ve^ CD25^+ve^ CD127^low/-^ Treg population in PTP1B and DUSP4 knockdown with IL-2 stimulation in PMOS subjects. FACS analysis showed an increase of CD4^+ve^ CD25^+ve^ CD127^low/-^ Treg population after PTP1B and DUSP4 (MKP-2) siRNA-based knockdown in PBMCs of PMOS (n=12) subjects. A: bar graph representation showing the gene expression of *PTP1B* and *DUSP4* across the PTP1B, DUSP4, and both silenced PBMCs. B: Dot plot and histogram representation of gating strategy with P1 shows lymphocyte population, P2 shows CD4-FITC^+ve^ population, P3 shows CD25-PE Cy7^+ve^ population and P4 shows CD127 ^low/-^ Treg population. Gating is based on histogram of unstained and stained population (FITC, PE Cy7 and Alexa Fluor 647 respectively). C: Dot plot representation of CD4-FITC^+ve^ CD25-PECy7^+ve^ CD127-Alexa Fluor 647^low/-^ (P4) Treg population of all treatment conditions including; untreated, IL-2 treated, control siRNA silenced, control siRNA silenced with IL-2, PTP1B siRNA silenced without IL-2 and with IL-2, DUSP4 siRNA silenced without IL-2 and with IL-2, and PTP1B+DUSP4 siRNA silenced without IL-2 and with IL-2. D: Dot and truncated violin plot representation of the percentage of CD4^+ve^ CD25^+ve^ CD127^low/-^ Treg population in lymphocyte events across all the treatments. Each dot represents an individual subject. One-way ANOVA was performed using GraphPad Prism 8 and p-value< 0.05 were considered as statistically significant.

#### A. No change in Treg population after IL-2 treatment (Figure 7D-1 and 7D-2)

The FACS analysis showed no change in CD4^+ve^ CD25^+ve^ CD127^low/-^ Treg population between untreated PBMCs and IL-2 stimulated PBMCs of PMOS subjects as shown in Figure 7D-1 and 7E. This finding again corroborates our earlier findings where IL-2 stimulation of PBMC from PMOS subjects did not cause increased FOXP3 expresssion due to inapt STAT5 phosphorylation^19^. We also observed no change in CD4^+ve^ CD25^+ve^ CD127^low/-^ Treg population between control silenced PBMCs and control silenced+IL-2 stimulated PBMCs.

#### B. No change in Treg population after individual silencing with IL-2 treatment (Figure 7D-3 and 7D-4)

The FACS analysis showed no significant alteration of CD4^+ve^ CD25^+ve^ CD127^low/-^ Treg population after silencing of *PTP1B* expression. Furthermore, even after addition of exogenous IL-2, there is no significant change in Treg population in the *PTP1B* silenced PBMCs (Figure 7D-3). Similarly, we also observed no significant alteration of CD4^+ve^ CD25^+ve^ CD127^low/-^ Treg population in *DUSP4* silenced PBMCs with and without IL-2 treatment.

#### C. Increased Treg population after double silencing with IL-2 treatment (Figure 7D-5)

Our further analysis revealed a significant increase of CD4^+ve^ CD25^+ve^ CD127^low/-^ Treg population in IL-2 stimulated *PTP1B* and *DUSP4* double silenced PBMCs, as compared to the control silenced with IL-2 treatment (p-value <0.05) (Figure 7D-5 and 7E). Thus, our results clearly demonstrate that women with PMOS exhibit a decrease in Treg due to increased phosphatase expression and activity. Treg frequency increases when the effects of these phosphatases (PTP1B and DUSP4) are blocked. This implies that the increased PTP1B and DUSP4 may have a role in lowering the frequency of Tregs in PMOS women.

## DISCUSSION

The host tolerance mechanism was most likely designed to maintain host homeostasis by coordinating the harmful burden of infections with the immune-driven resistance strategy of the tissue damage control system. It is possible to achieve this homeostasis through central and peripheral tolerance. Central tolerance occurs during the development of immune cells, while peripheral tolerance happens throughout life, regulating responses toward foreign antigens entering our bodies, thereby maintaining homeostasis. Regulatory T cells are key players in providing peripheral tolerance mechanisms, and studies have shown that the CD4^+^ CD25^+^ regulatory T cells in naïve mice show *in-vitro* suppressive action by suppressing the proliferation of other T cells ^34^. PMOS has been well-documented as a chronic inflammatory condition ^35,36^ and decreased peripheral Treg reported in PMOS^37^ suggests a reduced immune tolerance capability. Treg differentiation is masterminded by an IL-2-mediated JAK-STAT5 pathway with STAT5 having an essential role in Treg homeostasis^38^. Interestingly, no alteration in IL-2 levels is observed between normal and PMOS subjects in our study, suggesting that the defective STAT5 phosphorylation is independent of IL-2 status. STAT5 phosphorylation via JAK is a crucial step in STAT activation. Our findings of a reduced JAK2 in PMOS subjects could explain the reduced STAT5 phosphorylation in PMOS, thereby implicating it in the low frequency of Tregs. This is supported by the existence of an active IL-2-driven Jak-STAT5 signalling for FOXP3 maintenance in humans, which is sensitive to Jak inhibitor (AG-490)^39^. Findings of an increased PTP1B phosphatase could dephosphorylate the already reduced JAK2 levels, an established substrate of PTP1B^40^. Also, the overexpressed Jak 1 could be reined in by its dephosphorylation due to elevated TCPTP (Figure 2), whose established target is JAK1^41^.

Dephosphorylation of STATs is one of the main mechanisms crucial for controlling the strength of STAT5 signalling. Our study on leukocytes in PMOS focused on phosphatases regulating the STAT5 pathway for Treg differentiation, and we have found an upregulated gene expression of phosphatases; *PTP1B, TCPTP*, and *DUSP4* (Figure 2). In tandem with this, the PBMCs from PMOS women had a significantly increased phosphatase activity (Figure 5). This finding explains the dephosphorylated state of STAT5B in PBMCs from PMOS women reported earlier by us^19^.

Amongst the three phosphatases, our elevated PTP1B data corroborate with a similar study on breast cancer, which reported that PTP1B knockdown increases the tyrosine phosphorylation of STAT5, while its overexpression minimises phosphorylated STAT5 expression in response to prolactin stimulation ^42^. Testosterone has been shown to reduce CD4 T-cell differentiation by elevating PTP1B encoding gene *Ptpn1* expression via androgen receptor binding to a conserved region in intron 3 of *Ptpn1* ^43^. Our findings of an elevated DUSP4 in PMOS women hold important significance as Huang et al., have reported that in DUSP4 (-/-) mice, the DUSP4 (-/-) CD4 ^+^ T cells hyperproliferation is induced by IL2 signalling via elevated CD25 expression followed by STAT5 phosphorylation^44^.

Upon further classifying PMOS patients based on their phenotypes, it was found that DUSP4 was the only phosphatase that was upregulated in those patients with phenotype C, while patients with phenotype A had considerably increased expression of all three phosphatases (Figure 3). Given that phenotypes A and C share characteristics with hyperandrogenism and PCOM, the pathophysiology of hyperandrogenism and polycystic ovary morphology may be related to an increase in phosphatase expression. It’s interesting to note that our linear regression correlation analysis (Figure 4) revealed a significant association between elevated LH and testosterone levels with *DUSP4* only, suggesting that increased expression of DUSP4 has a crucial role in the pathogenesis of PMOS. The significant positive correlation between DUSP4 and testosterone seen in Figure 4 is supported by reports of Choi et al., who postulate a possible crosstalk between DUSP4 and androgen receptor^45^.

Both LH and hCG relay their action via luteinising hormone choriogonadotrophin receptor (LHCGR) activation. Luteinizing hormone is known to exert its action by phosphorylating MAPK, enabling its activation^46,47^. Interestingly, Luteinizing hormone receptor activation by hCG induces MAPK phosphatase-2 (MKP-2, *DUSP4*)^48^ suggesting that elevated LH in PMOS could also induce DUSP4. Our earlier reports of overexpressed p38 kinase in PMOS women^49^ and the fact that DUSP4 does not dephosphorylate p38MAPK while it dephosphorylates ERK^50^, suggests p38 MAPK’s possible role in triggering DUSP4 overexpression. This gains more traction from our findings of elevated STAT3 gene and protein expression and its increased phosphorylation status as a consequence of the overactivated p38 MAPK pathway^49^, since STAT3 is shown to positively regulate the transcriptional activity of DUSP4 Variant 1 via a STAT3 binding site in its promoter^51^.

The leukocytes contain a notable proportion of neutrophils (40% - 60%) as well as monocytes and other granular immune cells. such as eosinophils, and basophils. Hence, decoding whether the above phosphatases (PTP1B, TCPTP, and DUSP4) play a role in regulatory T cells is an important question. PBMCs also contain 70% - 90% of lymphocytes, 10% - 20% of monocytes, and 1% - 2% of dendritic cells. Again, the lymphocytes consist of T lymphocytes, B lymphocytes, and NK cells. Regulatory T cells are T cell subtypes that comprise of 1% - 4% of the total PBMC population, and 5% −10% of total CD4^+ve^ T cells. Our subsequent analysis of phosphatases on CD4^+ve^ CD25^+ve^ Treg cells confirmed an upregulation in the gene expression of *PTP1B* and *DUSP4* in PMOS women (Figure 6), whereas the difference in the expression of *PTP1B, TCPTP,* and *DUSP4* was insignificant among the CD4^−ve^ cells, and CD4^+ve^CD25^−ve^ T effector cells in the control and PMOS women. This evidence uncovered that the upregulation of PTP1B and DUSP4 may be involved in compromising CD4^+ve^ CD25^+ve^ Treg differentiation in PMOS women. JAK-STAT signalling is the major pathway involved in the regulation of the immune system^52^. Studies in different conditions established a significant paradoxical role of PTP1B in attenuating the JAK-STAT pathway by dephosphorylating JAK2^40,53^. A recent finding by Wiede et al 2022, revealed that in the intra-tumoral CD8^+^ cells, PTP1B induction limits its expansion and cytotoxicity, while its deletion enhances the activity and cytotoxicity of CD8+ T cells by increasing STAT5 signalling ^54^. Furthermore, in support of our result of elevated presence of DUSP4 in the CD4^+ve^ CD25^+ve^ Treg cells only in PMOS conditions, Huang CY et al revealed that the dual-specificity phosphatase, DUSP4, established a negative effect on IL-2 signalling by dephosphorylating pSTAT5 and hence, inhibits CD4 T cell proliferation ^44^. In addition to this, Hsiao et al reported that DUSP4 deficient T cells enhance in-vitro differentiation of iTreg while reducing polarization of Th17 cells ^33^, which corroborates our findings of low tregs^19^ with high DUSP4 expression. The insignificant difference in TCPTP in CD4^+ve^ CD25^+ve^ suggests that it may not have a role in STAT5 dephosphorylation in T cells. This is strengthened by the lack of support to TCPTP mediated STAT5 dephosphorylation in TCPTP-knockout mice^55^. Hence, this signifies that the overexpression of PTP1B and DUSP4 in the CD4^+ve^ CD25^+ve^ Treg cells of PMOS patients would have a compounding impact in the dephosphorylation of peripheral STAT5 leading to lowered Treg frequency as reported earlier by us^19^.

The significant increase of CD4^+ve^ CD25^+ve^ CD127^low/-^ Treg population observed in IL-2 stimulated PBMCs following *PTP1B* and *DUSP4* silencing, compared with control siRNA treated IL-2 stimulated cells, highlights that targeting phosphatases may represent a promising strategy to enhance Treg levels. This is reinforced by a recent study of use of PTPN1/PTPN2 inhibitor “ABBV-CLS-484”, which releases mighty anti-tumour immunity by improving JAK-STAT signalling and correcting T-cell dysfunction^56^. Together, these findings position PTP1B and DUSP4 as promising, druggable immunotherapeutic targets.

The development of selective phosphatase inhibitors will be essential for translating these insights into strategies to restore Treg populations and re-establish immune tolerance across disease settings.

## Conclusion

In conclusion, the present study identifies PTP1B and DUSP4 as key negative regulators of IL-2–driven Treg expansion in PMOS, underscoring the potential of phosphatase targeting to fine-tune immune homeostasis (Graphical Abstract). By sustaining IL-2/STAT5 signaling and enhancing Treg differentiation, modulation of these phosphatases emerges as a rational strategy to restore immune tolerance in conditions marked by Treg insufficiency. A key limitation of this study is the lack of *<u>in vivo</u>* validation, owing to the unavailability of specific inhibitors for these phosphatases. Future work should focus on developing selective inhibitors or employing transient knockdown approaches to determine whether functional Treg expansion can be reliably achieved *<u>in vivo</u>*. Such efforts will be critical for translating these mechanistic insights into clinically relevant immunomodulatory therapies. Collectively, our findings advocate a shift beyond PD-1–centric immunotherapy toward targeting phosphatases as promising and druggable regulators of T-cell dysfunction.

## MATERIAL AND METHODS

### Patient enrolment and sample collection

This study is a retrospective study involving 102 control and 102 PMOS female donors of reproductive age (18-40 years). On the early follicular phase (day 2-5) of each subject’s menstrual cycle, 10 mL of peripheral blood was drawn. Samples were collected from KJK hospital and SAMAD IVF clinic located in the extreme southern part (Kerala) of India. Clinical diagnosis of PMOS patients was done based on the Rotterdam Consensus criteria (The Rotterdam European Society for Human Reproduction and Embryology (ESHRE)/ American Society for Reproductive Medicine (ASRM)-sponsored PMOS Consensus Workshop Group, 2004). Control subjects were selected as per the criteria of regular menstrual cycles, without any history of metabolic/hormonal imbalance, and no fertility problems. Subjects with other androgen disorders, autoimmune disorders, and infections within 3 weeks were excluded from our study.

#### Ethics statement

The Institute Ethical Review Board of Rajiv Gandhi Centre for Biotechnology (RGCB) gave ethical approval for this work under approval numbers IHEC/01/2019/07 and IHEC/01/2015/09. Blood samples were collected from control and PMOS/PCOS individuals after informed consent was obtained according to the Declaration of Helsinki.

### Hormone profiling by ELISA

The plasma levels of Follicle Stimulating Hormone (FSH), Luteinizing Hormone (LH), Testosterone, and Dihydrotestosterone (DHT) were estimated in all subjects by using human FSH (# 500710), LH (#500720), and Testosterone (#582701) ELISA kits (Cayman chemicals, USA) and DHT ELISA kit (MBS2000056, MyBiosource, USA).

### IL-2 ELISA

The plasma IL-2 levels were assessed in 37 control and 37 women diagnosed with PMOS by using the BD Opt EIA Human IL-2 Elisa kit II (catalogue No. 550611) as per manufacturer’s protocol.

### Quantitative real-time PCR (qRT-PCR) in leukocyte RNA

Isolation of total leukocyte RNA was done from 1 mL of peripheral blood of both control (n=102) and PMOS (n=102) subjects using miRNeasy mini kit (#217004, Qiagen, Germany) as per the manufacturer’s instructions. Using SuperScript^TM^ VILO^TM^ cDNA Synthesis Kit (11754-050, Invitrogen, USA), cDNA was synthesized, and then a qRT-PCR was carried out with Power SYBR^®^ Green (4367659, Life Technology, USA) on a 7900HT Fast Real-Time PCR system (Applied Biosystems). NCBI PRIMER BLAST was used to design oligonucleotide primers for real-time analysis (Table S1) and the designed primers were obtained from Eurofins, India and Sigma Aldrich. The relative expression of all the genes were calculated with ΔΔCt method. 18S rRNA was used as endogenous control. The fold change (FC) was calculated and represented as 2^−ΔCt^. The student’s t-test was conducted and graphs were created with GraphPad Prism 8. Statistical significance was attained when the p-value<0.05.

### Protein lysate extraction from PBMCs and phosphatase assay

PBMCs were isolated using the conventional Ficoll-Histopaque density gradient centrifugation method with 8 mL of whole blood^57^. Isolated PBMCs were incubated for 30 minutes at 4°C with 200μl of protein extraction buffer (50 mM HEPES (4-(2-hydroxyethyl)-1-piperazineethanesulfonic acid) (pH 7.4) containing 0.5% TritonX-100, 10% glycerol, and 1X protease inhibitor cocktail). The protein was extracted from the supernatant and was used immediately for the phosphatase assay. The levels of phosphatases in PBMCs of control (n=20) and PMOS (n=23) subjects were measured using a RediPlate 96 EnzChek® Tyrosine Phosphatase Assay Kit (R22067, Thermo Fisher Scientific, MA, USA) following the kit protocol. This is a fluorescence-based microplate assay that detects phosphatases (PTPase) in the sample by detecting the specific substrate i.e., DiFMUP (6,8-difluoro-4-methyl umbelliferyl phosphate), which gets dephosphorylated by the action of phosphatases in the sample, yielding a reaction product, DiFMU (6,8-difluoro-4-methyl umbelliferyl). The DiFMU exhibits fluorescence at excitation/emission maxima of 358nm/452nm and the fluorescence reading was obtained using Varioskan LUX Multimode Microplate Reader (Thermo Fisher Scientific). The phosphatase level was normalized to the total protein level of the sample. Graphs were created using GraphPad Prism 8, and a t-test was run to ascertain the statistical significance; a p-value<0.05 was considered statistically significant.

### FACS analysis and qRT-PCR of magnetically isolated regulatory T cells from the PBMCs

1 to 2 × 10^6^ PBMCs were isolated from both control (n=11) and PMOS (n=13) subjects. PBMCs count were determined with Tali® Image-Based Cytometer (Invitrogen). Regulatory T cells were isolated from PBMCs of both control and PMOS subjects using Dynabeads™ Regulatory CD4^+^ CD25^+^ T Cell Kit (Catalog no-11363D, Thermo Fisher Scientific, USA) in line with the manufacturer’s guidelines. As a result of magnetic separation, three types of cells were separated: CD4^−ve^ cells (non-CD4 cells), CD4^+ve^ CD25^−ve^ (effector) T cells, and CD4^+ve^ CD25^+ve^ regulatory T cells from the collected PBMCs of both control and patients. To validate the accuracy of magnetically isolated CD4^+ve^ CD25^+ve^ regulatory T cells, FACS analysis was performed using isolated normal PBMCs from a control female and magnetically separated CD4^+ve^ CD25^+ve^ regulatory T cells. The normal PBMCs were classified as Unstained normal PBMCs and used for gating the autofluorescence-negative and positive staining. Also, another set of CD4^+ve^ CD25^+ve^ regulatory T cells were classified as Unstained-Magnetically separated cells and used to check the accuracy of magnetically isolated cells. The magnetically separated cells were incubated with antihuman CD3 APC-H7 (item no. 560176), antihuman CD4 Per CP Cy5.5 (item no. 560650), and antihuman CD25 PE (item no. 555432) according to the manufacturer’s instructions (BD Biosciences, USA). FSC and SSC parameters were set to the optimal threshold and 20,000 lymphocyte events were gated as P1(lymphocyte) population. From the lymphocytes, CD3 APC-H7^+ve^ T cells were gated as the P2 population. Subsequently, double positive for both CD4 Per CP Cy5.5^+ve^ and CD25 PE^+ve^ Treg cells were gated as Q2 population from the CD3^+ve^ T cells through BD FACS DIVA software.

The total RNA was extracted from each cell type and finally to determine the expression levels of target genes, qRT-PCR was performed. The computation of the FC was represented as 2^−ΔCt^. To determine the statistical significance, the student’s t-test was computed and graphs were created with GraphPad Prism 8. Statistical significance was defined with a p-value<0.05.

### RNA interference and FACS analysis

5×10^5^ PBMCs were isolated from PMOS (n=12) patients and seeded on a 24-well plate (Thermo Fisher Scientific) and cultured in RPMI 1640 (ATCC) complete media supplemented with 10% FBS (#10437-028, Invitrogen, USA) and 1X antibiotic-antimycotic cocktail (#15240-062, Gibco, Thermo Fisher Scientific, MA, USA) and incubated at 37°C and 5% CO2 for 24 hours. To knockdown PTP1B and DUSP4 (MKP-2) in PBMCs, the PBMCs were transfected twice with 100 nmol concentration of small interfering RNA (siRNA) specific for human PTP1B (Sc-36328) and/or DUSP4/MKP-2(MAPK phosphatase-2) (sc-38998) or control siRNA (non-targeted, sc-37007) (Santa Cruz Biotechnology) using Lipofectamine 3000 (Cat #L3000-015, Life Technologies, USA) in Opti-MEM^TM^ medium (Cat #31985070, Thermo Fisher Scientific, USA). After 24 hours of siRNA treatment, the cells were stimulated with 300IU of recombinant IL-2 (rIL-2) (#130-097-743, Miltenyi Biotech, Germany). Finally, after treatment, the cells were collected for subsequent Fluorescence-activated cell sorting (FACS) analysis and expression analysis via qRT-PCR. For FACS analysis the cells were harvested and incubated for 40 minutes with 0.1% BSA-PBS and Human Regulatory T Cell Cocktail (#560249, BD Pharmingen^TM^, USA), having antihuman CD4 fluorescein isothiocyanate, antihuman CD25 PE cy7, and antihuman CD127 Alexa fluor 647. After incubation the cells were washed twice and acquired using BD FACS Aria III flow cytometer. Analysis of fluorescence intensity was performed and a total of 20,000 lymphocyte events (P1) were gated through forward scatter vs side scatter from the total population, followed by gating of the CD4^+ve^ population (P2). Further, gating was performed based on CD25^+ve^ (P3) and then the CD127^low/-^ population (P4) was selected to analyse CD4^+^ CD25^+^ CD127^low/-^ Treg cells.

### Statistical Analysis

All the graphs were created using GraphPad Prism 8 and statistical significance was defined with a p-value<0.05. The clinical and biochemical variables were represented as Mean±SEM. Unpaired Student’s T-test was performed to calculate the significance level of all the biochemical parameters, IL-2, enzyme activity of phosphatases, and real-time gene expression of *JAKs* as well as phosphatase genes in our recruited human samples. One-way ANOVA with Dunnett’s multiple comparison test was conducted for the real-time gene expression comparison of *JAKs* and phosphatases between four different phenotypic PMOS subgroups. One-way ANOVA with Tukey’s multiple comparison test was performed to check the significance difference between various groups in siRNA-based transfection experiment. Linear regression correlation analysis was conducted to check the correlation of hyperandrogenism with other biochemical parameters, including LH, LH/FSH, Testosterone, DHT, and BMI. The Excel data analysis tool was used to calculate the statistical calculations and data analysis for all the experiments.

## Data availability statement

The data that support the findings of this study are available in the Main Text and in the online Supplementary material. Certain data underlying Table 1 are not publicly available due to privacy or ethical restrictions.

## Acknowledgements

We acknowledge the support extended by KJK Hospital and SAMAD IVF Hospital staff for patient recruitment and data management. The normal and PMOS volunteers are also acknowledged. This work was funded by an ICMR Grant (No.5/10/FR/50/2020-RBMCH) and Rajiv Gandhi Centre for Biotechnology Core Funds from the Department of Biotechnology to M.L. ML is currently supported as an Emeritus Scientist by ICMR (HRD/Head/IES/2024/9). L.P.P. was supported by a Research Fellowship from the University Grants Commission; No.435/(CSIR-UGC NET DEC.2018). B.S.J. was supported by a research fellowship from the Department of Science and Technology (DST/INSPIRE fellowship/2015/1F150361).

## Author contributions

M.L. contributed to project conception, study design, experiment supervision, data interpretation, and manuscript review. M.L. and L.P.P. designed the Graphical Abstract. L.P.P. and B.S.J. collected and processed the samples, and executed the experiment for Figure 2. L.P.P. executed the experiments for Figures 1 and 3-7, wrote the first draft of the manuscript, analysed and interpreted all the data for Figures 1-7. The diagnosis and recruitment of PMOS patients was done by K.J.K. and S.M.P. All authors were involved in the discussions of the results.

## List of Supplementary Material

Supplementary Table S1

Fig. S1

