## Supplementary material for "Targeting PTP1B and DUSP4 phosphatases to Boost Tregs: A Novel Therapy for Polyendocrine Metabolic Ovarian Syndrome (PMOS) Immune Dysfunction": Figure S1

#### Supplementary Figure: Figure S1

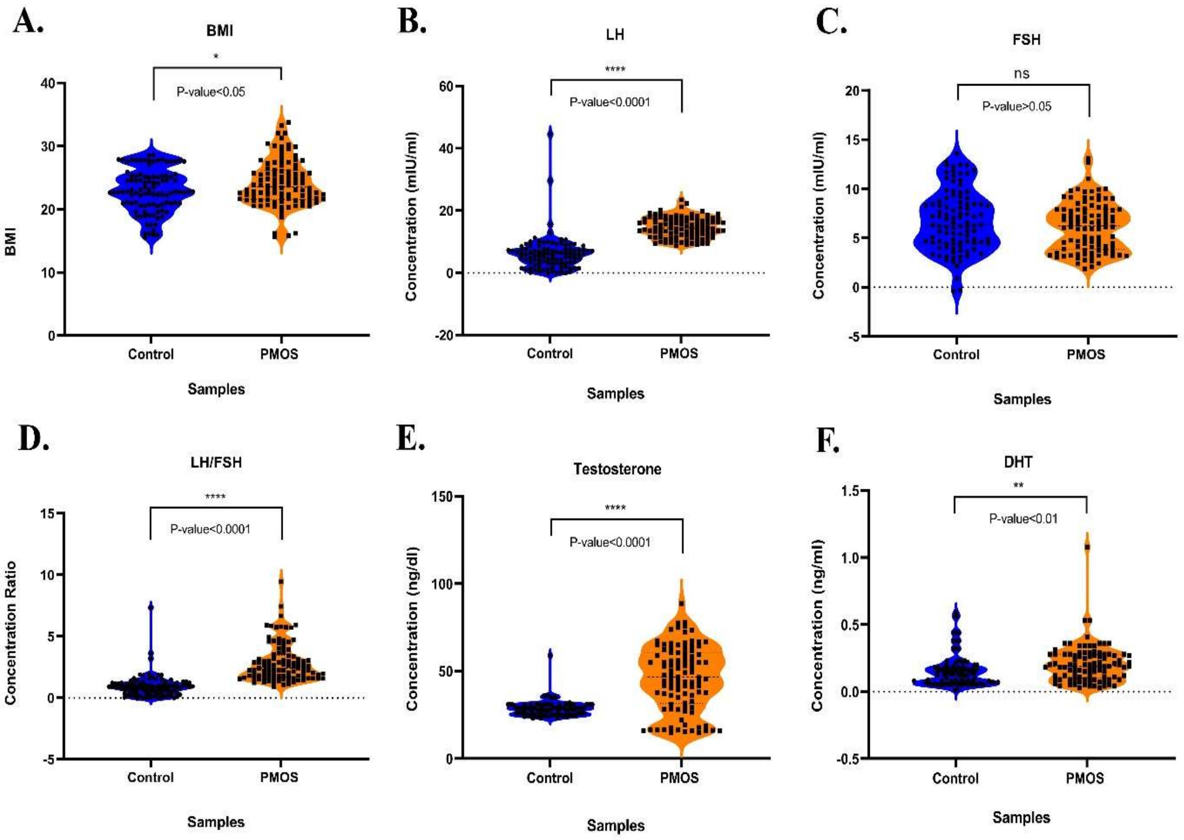


### Figure S1

***Plasma level of various hormonal parameters in control and PMOS subjects.*** *Plasma level expression of various hormonal parameters in 102 control and 102 PMOS subjects. A-F: Dot and violin plot graphical representation of A: BMI and B-F -plasma level expression of B: LH, C: FSH, D: LH/FSH, E: Testosterone, and F: DHT in control and PMOS subjects of our study group. Each dot inside the violin plot represents an individual subject. Unpaired student's T-tests were performed using GraphPad Prism 8 and p-value< 0.05 were set to be significant.*
