## Supplementary material for "Targeting PTP1B and DUSP4 phosphatases to Boost Tregs: A Novel Therapy for Polyendocrine Metabolic Ovarian Syndrome (PMOS) Immune Dysfunction": Table S1

**Supplemental Table S1: Primer Details**

| ***Gene*** | ***Forward primer*** | ***Reverse primer*** |
|---|---|---|
| *h18S rRNA* | *CGGAACTGAGGCCATGATTA* | *CTTTCGCTCTGGTCCGTCTT* |
| *hJAK1* | GAGACAGGTCTCCCACAAACAC | GTGGTAAGGACATCGCTTTTCCG |
| *hJAK2* | CACCAGCGGAATTTATGCGT | GGCATCCATCTGGTCTTGGT |
| *hPTP1B* | *AGACGTCAGTCCCTTTGACC* | *TGACCGCATGTGTTAGGCAA* |
| *hDUSP4* | *CATAGCAGATCGCCCAGGAG* | *CTCATAGCCGCCTTGAGACT* |
| *hTCPTP* | *AAGCCCACTCCGGAAACTAAA* | *AAACAAACAACTGTGAGGCAATCTA* |
